# Protocol for a Comprehensive Analysis of Hepatitis C Risk Factors among the Rohingya Population in Camp Settings in Cox’s Bazar: A Mixed-Method Study

**DOI:** 10.1101/2025.04.02.25325119

**Authors:** Charls Erik Halder, Md Abeed Hasan, Hamim Tassdik, James Charles Okello, S M Niaz Mowla, Aarti Shrikrishana Singh, Partha Pratim Das, Md Arafat Hossen

## Abstract

**Background:** The hepatitis C virus (HCV)—a communicable disease with global health concerns—affects approximately 20% of the Rohingya refugee population in Cox’s Bazar, Bangladesh, which is higher than in its endemic countries and host population. While researchers have yet to determine specific transmission risk factors for this vulnerable, underserved, and overlooked cohort, the HCV seroprevalence is significantly high. Thus, this study will assess socio-demographic, medical, traditional, and behavioral factors contributing to HCV risk among the Rohingya refugee population to provide the basis for future prevention efforts.

**Methods:** A mixed-method study comprised of both qualitative and quantitative approaches will be employed. For phase one, a case-control study of 900 HCV positive cases and 900 matched HCV negative controls will be conducted to determine associations with risk factors through semi-structured questionnaires. For phase two, focus group discussions (n=12) and key informant interviews (n=30) with caregivers, healthcare providers, and community members will help explain and expand on quantitative findings. Analyses will include logistic regression (quantitative) and thematic analysis (qualitative).

**Expected Outcomes:** The study aims to identify key risk factors including unsafe medical practices, traditional procedures, personal hygiene behaviors, and sexual practices—and assess their association with HCV seropositivity. Findings will support recommendations for evidence-based prevention and policy and community initiatives to reduce HCV transmission among refugee populations.

**Ethics & Dissemination:** Ethics approval was obtained from Cox’s Bazar Medical College Hospital. Study findings will be disseminated via publication in a peer-reviewed journal and workshops with stakeholders to guide public health response.

**Author summary:** Hepatitis C is an overlooked public health issue in the refugee setting. We intend to determine the risk factors for the elevated rates of Hepatitis C among the Rohingya refugees living in the camps of Cox’s Bazar in Bangladesh. Using a mixed-method approach, we will use quantitative epidemiological analysis of medical information and risk behavior collected from 1,800 refugees and qualitative information collected from focus groups with (medical) community stakeholders. The expected results will allow for implementation of a culturally specific motivation for prevention and evidence-based policy for reduced Hepatitis C transmission in refugee settings. A transdisciplinary project with researchers/academics from IOM, Save the Children, WHO and local/international NGOs.

## Introduction

Hepatitis C, an inflammation of the liver caused by the Hepatitis C virus (HCV), remains a global health threat, causing 242,000 deaths annually (WHO, 2024a). The hepatitis C virus 1 can cause both acute and chronic hepatitis C and may result in lifelong diseases, especially liver cirrhosis and hepatocellular carcinoma. Globally, around 50 million people are suffering from chronic hepatitis C with one_1_ million new cases every year (WHO, 2024b). Globally, only 36.4% (as of 2022) of people living with hepatitis C are diagnosed, and only 20% receive treatment, against a target of 90% to be tested and treated by 2030 (WHO, 2024b). Access to affordable medicines, policy and programmatic barriers and lack of priority in funding Hepatitis C hinder access to treatment (WHO, 2024b). Refugees and displaced people are vulnerable to Hepatitis C due to their limited access to healthcare and information on its transmission and prevention, traditional practices, sexual violence and increased risky behaviors caused by isolation and stress (Ayele et al., 2020).

While previous studies reported a high prevalence of Hepatitis C among the Rohingya population in camp settings in Cox’s Bazar, ranging from 8% among the pregnant Rohingya women (Ali et al., 2022) to 13.2% among the Rohingya population (Al Mahtab et al., 2019), a recent study conducted by Médecins Sans Frontières (MSF) found that an estimated 20% of the Rohingya population in camp settings in Cox’s Bazar have active hepatitis C (Schramm & Ashakin, 2023). In contrast, the prevalence reported in other high-burden settings are 5.9% in Gabon, 3.6% in Burundi, 3.1% in Ukraine and 2.9% in Russia (CDC, 2024). Compared to other refugees globally, data from the study suggest HCV prevalence is three-fold higher than the highest estimate, 5.6%, among refugees and immigrant populations originating from high and intermediate HCV endemic countries (Ali et al., 2022b). Prevalence of Hepatitis C among the general population in Bangladesh is between 0.2 - 1% (WHO, 2024c) and in Myanmar is 2.7% (Scott et al., 2021). This demonstrates an extreme high prevalence of hepatitis C among the Rohingya population in camp settings in Cox’s Bazar which is alarmingly higher than general population in their residing country as well as their country of origin, general refugee settings and any other high burden settings studied. Therefore, it is imperative to understand the underlying factors that are contributing to the high transmission of Hepatitis C among the Rohingya population in camp settings in Cox’s Bazar. Literature highlights the mental health impacts of Hepatitis C infection, including increased rates of depression and anxiety, particularly in populations with high co-infection rates (Faccioli et al., 2021).

Empirically, Hepatitis C mainly spreads through contact with blood and the common risk factors include sharing needles or syringes or unsafe medical procedures such as blood transfusions with unscreened blood products (WHO, 2024a). Studies reported a range of other risk factors in different settings, for instance, circumcision by informal healthcare providers in Upper Egypt and Nile Delta (Medhat et al., 2002), injection drug use in Mexico and Georgia (Belaunzarán-Zamudio et al., 2017, Hagan et al., 2019), tattooing in Mexico (Belaunzarán-Zamudio et al., 2017), inadequate infection control in hospital setting in Georgia and having multiple sex partners (Hagan et al., 2019). The study of MSF identified that the factors associated with HCV seroprevalence include being female, age group 18 – 25 years, those who have history of surgery and those who reported medical injection (Schramm & Ashakin, 2023). However, there is no concrete evidence regarding which particulars factors or concerns in this context that are causing the transmission—for example, why the transmission rate is higher in women, or among the people who have had surgery or medical injections in the past, or, what kind of surgery or medical procedures are related to the transmission. Considering the existing refugee setting and previous history of the population in Myanmar, experts suggested the need to explore the possibility of different context specific factors, specially, exposure of the refugees to unsafe medical procedures in Myanmar or in camp setting, exposure to unsterile medical equipment within the camps, circumcision practices with unsterile instruments, intravenous drug use, exposure to contaminated personal care items (e.g. shaving instruments in barbershops), traditional cosmetic procedures (ear and nose piercing)(Ali et al., 2022, Schramm & Ashakin, 2023, Al Mahtab et al., 2019). Sexually transmitted infections are prevalent in Myanmar, with Rohingya girls and women facing heightened risk due to gender-based violence and exploitation during humanitarian crises. Increased reports of high-risk sexual behavior within the Rohingya community highlight the need for a focused study on Hepatitis C transmission through sexual contact in this population (Hossain, Sultana, and Mazumder, 2018).

MSF estimated that around 85,000 adults in the Rohingya refugee camps are already living with chronic Hepatitis C (Schramm & Ashakin, 2023). While prioritizing the treatment of this population is a priority, there is also critical need to prevent further transmission of Hepatitis C. This would require targeted interventions and extensive risk communication and community engagement centered around the prevalent risk factors. Therefore, it is crucial to comprehensively map out all potential risk factors and gain an in-depth understanding of why and how the factors are contributing to disease transmission in this particular setting.

### Research Questions

In the context of Rohingya refugee camps in Cox’s Bazar, Bangladesh:

1. What are the socio-demographic and medical history factors contributing to the high prevalence of Hepatitis C in the Rohingya camps, considering age groups and gender?
2. How do traditional and personal practices in the Rohingya context, including sexual activities /practices, contribute to the high prevalence of Hepatitis C?
3. What is the level of knowledge, attitudes, and practices (KAP) related to Hepatitis C preventive measures among the Rohingya population, and how is it associated with high disease transmission?
4. Why and how do the identified risk factors contribute to the high prevalence of Hepatitis C in the Rohingya refugee context.

## Materials and methods

### Study design

This will be a mixed-method study comprised of both qualitative and quantitative approaches aimed to explore and explain the risk factors associated with hepatitis C transmission in the Rohingya refugee camps in Cox’s Bazar. The study will be implemented during the period from 1st December 2024 to 30th April 2025. The study will have an explanatory sequential design (Wipulanusat et al., 2020). It consists of two phases:

- Phase 1: Quantitative (Case-Control Study): Intended to identify the risk factors associated with Hepatitis C transmission.
- Phase 2: Qualitative (Focus Group Discussions and Key Informant Interviews): Aims to elucidate the quantitative results and offer enhanced understanding of the risk variables.

In the first phase a case-control study will be implemented to identify the risk factors associated with Hepatitis C transmission among the Rohingya population in camp settings in Cox’s Bazar. Cases that are seropositive for Hepatitis C virus (HCV) and have detectable HCV Ribonucleic Acid (RNA), along with Rapid Diagnostic Test (RDT)-negative controls, will be selected from the Early Warning, Alert and Response System (EWARS) database, ensuring the inclusion of all age groups and maintaining. Recognizing the potential lack of formal medical records in the refugee camp setting, we will combine the available documents, such as any existing records of blood transfusions, surgical procedures, and hospitalizations, with self-reported histories from participants to triangulate and validate the medical history-related factors contributing to hepatitis C disease transmission.

In the second phase a qualitative study will be conducted to further understand and explain how the identified risk factors are contributing to the disease transmission in the refugee setting, including potential TB-HIV and HIV-HCV coinfections, and to generate evidence and recommendations for targeted intervention for disease prevention. Ethical considerations are detailed in the “Ethical Consideration” section of this document.

### Study Setting

The study will be implemented in Rohingya refugee camps in Ukhiya and Teknaf in Cox’s Bazar. Rohingya population staying in the 33 refugee camps and in the 2 registered camps will be the study population. The study will include participants from different age groups over 18 years, ensuring a representative sample. Age-specific questions will be included to capture relevant data for different cohorts. Gender balance will be maintained among cases and controls. The Rohingya population in camp settings in Cox’s Bazar are having a high prevalence of active hepatitis C impacting their lifespan, wellbeing and quality of life. Therefore, this setting has been selected for this study to get an in-depth idea of the risk factors associated with Hepatitis C high prevalence. In the qualitative phase, this study will explore not only the contextual factors contributing to these risks but also the socio-cultural practices and barriers to prevention that are unique to this population.

### Sampling Technique

For this study, a census sampling technique will be employed for the case group, whereby all 900 individuals enrolled in the cohort from IOM, WHO, and Save the Children who have been diagnosed with Hepatitis C will be included. This approach ensures comprehensive coverage of all eligible cases, maximizing the study’s power and providing a thorough analysis of the risk factors associated with Hepatitis C transmission in the refugee population. Census sampling is particularly beneficial in ensuring that no variation among cases is missed, providing full representativeness of the population studied (Rothman, Greenland, & Lash, 2008).

For the control group, matched random sampling will be used. Controls will be matched to cases based on key demographic criteria, including age, sex, and ethnicity. An age-sex distribution table will be created to ensure demographic parity between the two groups. Controls will be randomly selected from patients who tested negative for Hepatitis C (via RDT) at WHO sentinel sites, ensuring comparability between cases and controls while minimizing potential selection bias. This method follows established practices in epidemiological research to ensure the validity of control selection and reduce confounding (Schulz & Grimes, 2002).

## Phase 1: Quantitative (Case-Control Study)

### Study Population and Sampling

#### Cases

- A total of 900 individuals diagnosed with Hepatitis C (HCV-positive) will be selected.
- Inclusion criteria: Positive HCV antibodies confirmed by GeneXpert RNA testing. Cases will be identified through the EWARS database.
  ∘ Age 18 years or above

#### Controls

- Matched individuals (HCV-negative) selected based on:
  ∘ Same sex
  ∘ Age ±5 years
  ∘ Rohingya ethnicity
- Controls will be identified from WHO sentinel sites and hospital registers.

#### Selection Process

- Cases: Identified from the EWARS database, participants will be approached during routine follow-up visits for Hepatitis C treatment.
- Controls: Selected using a matched random sampling technique to ensure demographic parity with cases.

The case-control study is intended to assess the risk factors associated with Hepatitis C seropositivity by comparing two groups: cases, who have tested positive for HCV antibodies and have detectable HCV RNA, and controls, who have tested negative for Hepatitis C.

#### Case

900 Hepatitis C patients who are enrolled for treatment in International Organization for Migration (IOM) and Save the Children International (SCI) health facilities at Camp 2W and Camp 21 will be selected as cases for the study. These patients have positive serology test results for HCV antibody (detected by Rapid Diagnostic Tests) and detectable HCV RNA identified by GeneXpert confirmation. The patients will be identified from the EWARS database maintained by World Health Organization (WHO). The cases will be requested to participate in an enumerator-administered survey using a semi-structured questionnaire (see SI1 File) when they will attend the Hepatitis C clinics for treatment follow-ups.

#### Control

For each case a control will be recruited with matching criteria which include a) same sex, b) Rohingya ethnicity and c) within same age range (± 5 years). Thus, 900 controls will be recruited for the study from the patients attending the selected facilities and having negative RDT test results for Hepatitis C during their screening procedure. The control will also be identified from the EWARS database (with negative results) of WHO sentinel sites or hospital register (see Figure 1). To ensure a valid comparison, controls will be matched with cases based on key demographic criteria, including age, sex, and Rohingya ethnicity. An age-sex distribution table will be created to facilitate the selection of controls, ensuring they mirror the demographic characteristics of the case group. Controls will then be selected randomly from this balanced demographic pool, ensuring robust comparison between those with active Hepatitis C infection and those without any evidence of infection (Rothman, Greenland, & Lash, 2008). Similar to cases, the enumerators will request the controls to participate in an enumerator-administered survey using a semi-structured questionnaire (see SI1 File) on the same day as surveying the matching case.

**Figure 1.**
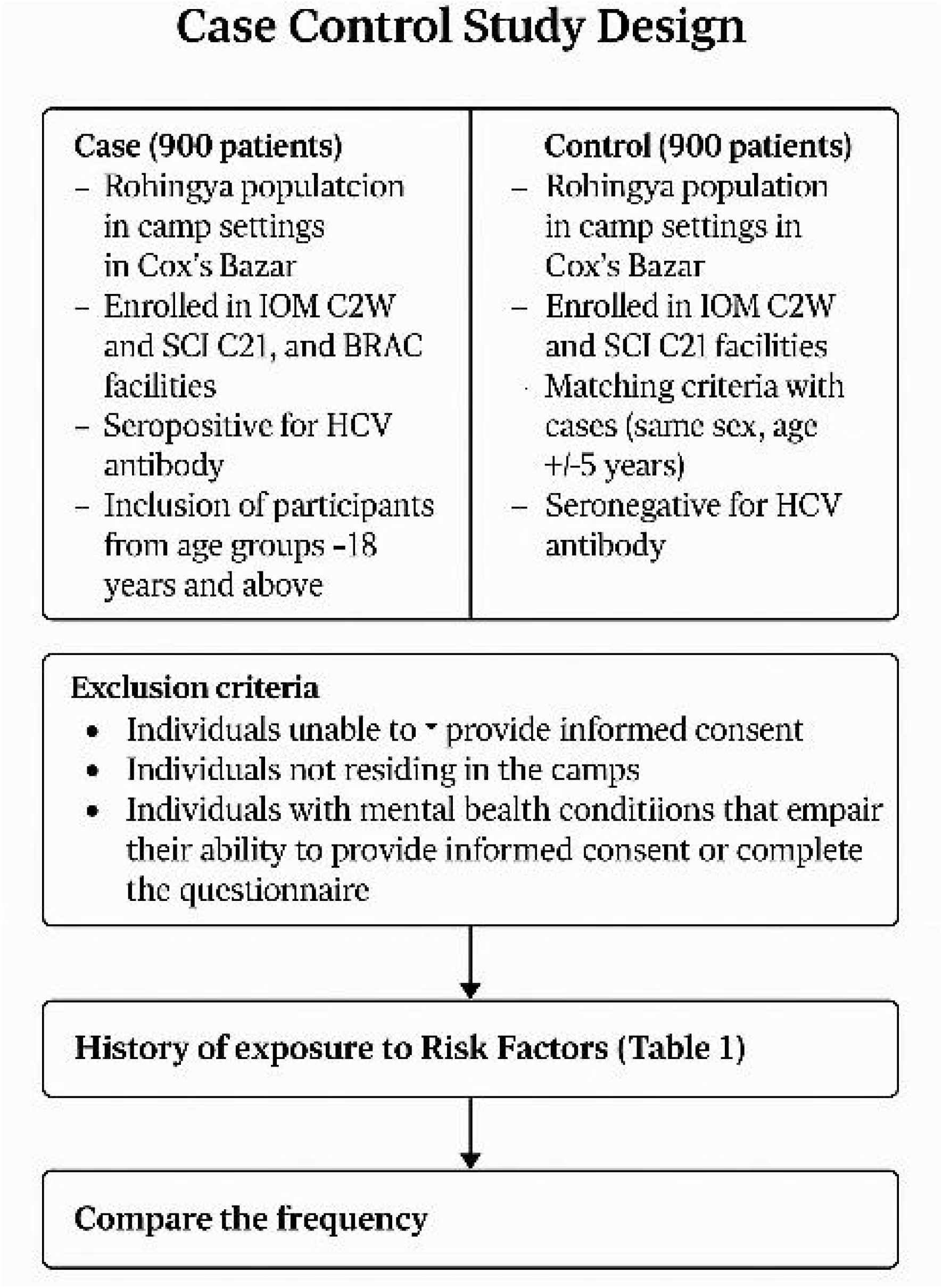
Case-control study design outlining the selection of cases and controls, exclusion criteria, assessment of exposure history, and comparison of risk factor frequencies.

### Data Collection

- **Tool:** Semi-structured questionnaires covering socio-demographics, medical history, high-risk behaviors, and Knowledge, Attitude, and Practice (KAP) adapted from Nawaz et al. (2018).
- **Method:** Enumerator-administered surveys using Kobo Toolbox.
- **Personnel:** 10 trained medical officers under the supervision of research assistants and the research lead.

Each of the cases and controls will be surveyed by an enumerator using a semi-structured questionnaire (see SI1 File) after obtaining informed written consent. The questionnaire form will be deployed through the Kobo toolbox for data collection. The questionnaire is developed in line with the research objectives and all potential risks identified from existing literature including socio-demographic, medical history, traditional practices, high-risk behaviours and sexual practices. The questionnaire also includes a knowledge, attitude and practice survey adapted from existing literature (Nawaz et al., 2018). 10 medical officers will be trained and deployed under the direct supervision of the research assistants and research lead for administration of the survey questionnaire and data submission using the Kobo toolbox.

**Table 1:**
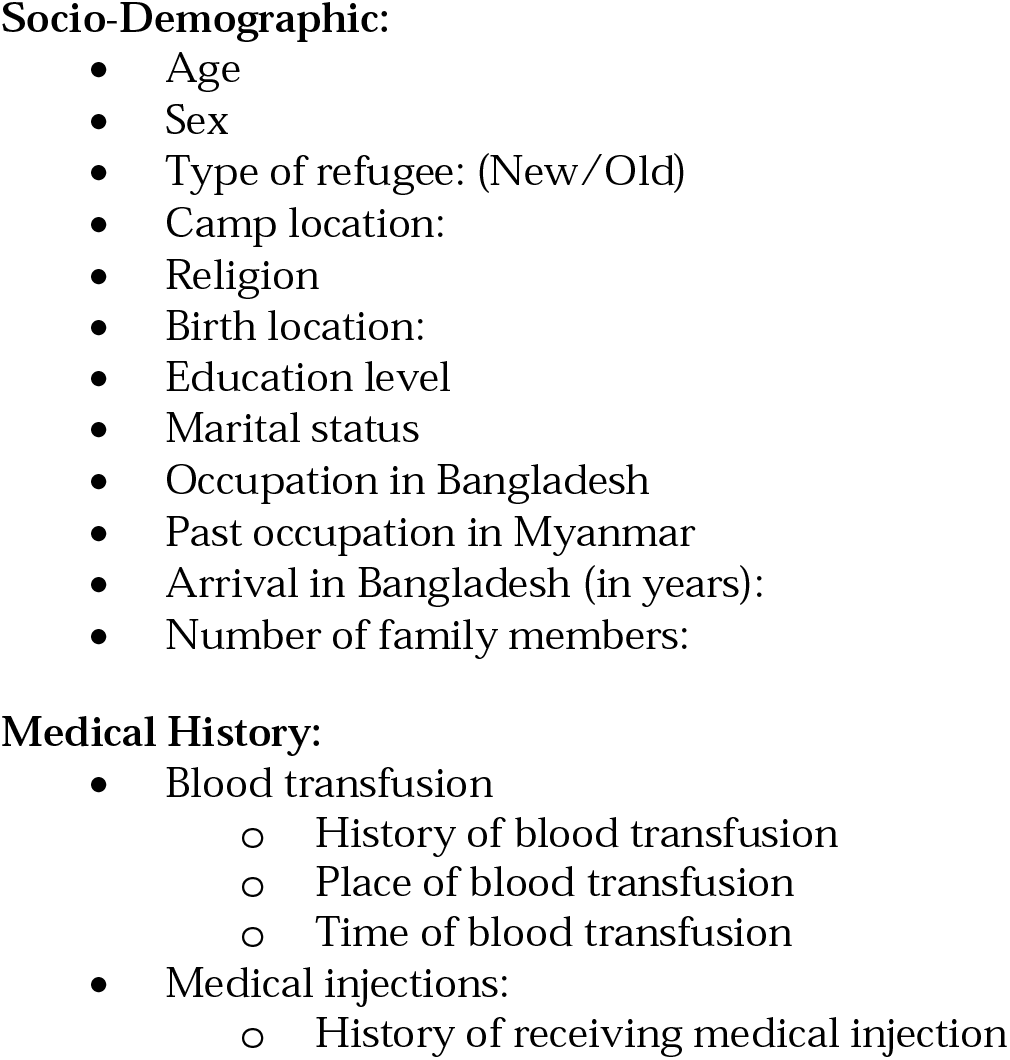
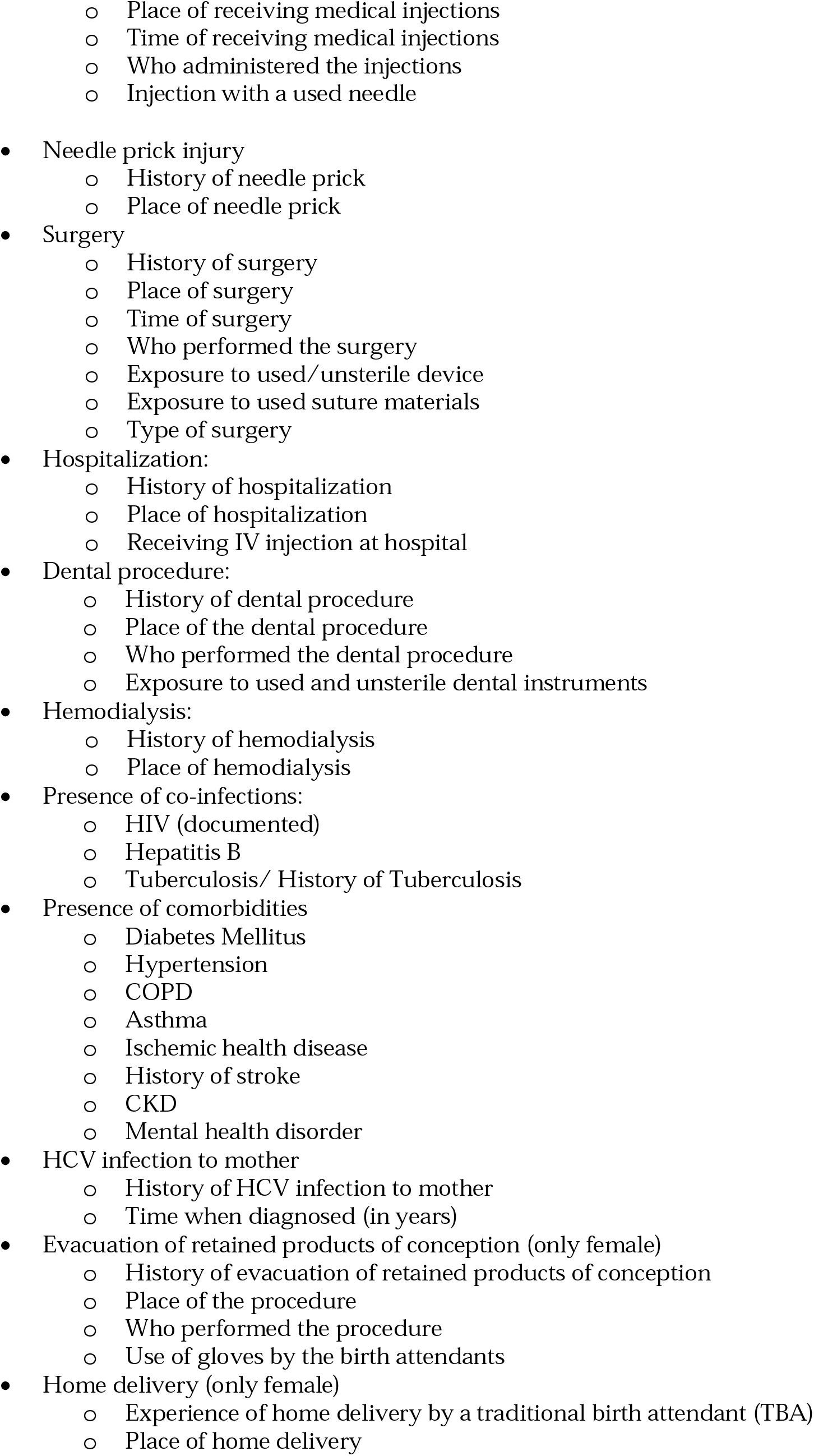
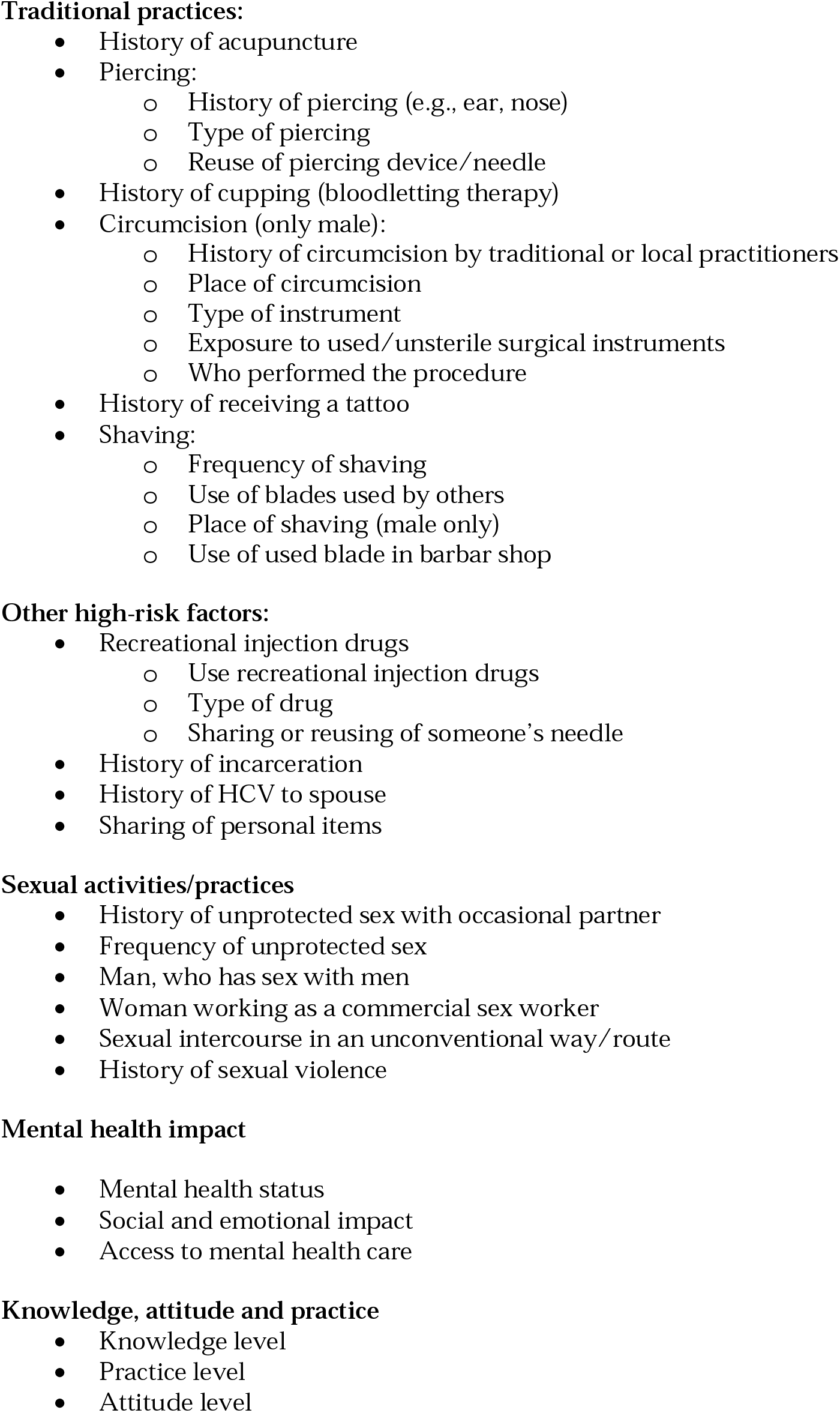
Independent variables – the potential risk factors for Hepatitis C Transmission.

## Phase 2: Qualitative (FGDs and KIIs)

The qualitative study aims to explain the findings of the case control study, including the nature of the risk factors and detailed description of risk factors, why the factors are prevalent among the Rohingya population in camp settings in Cox’s Bazar, how the factors contributing to disease transmission, and what can be done to prevent these risk factors with consideration local customs and context.

### Study Population and Sampling

- Focus Group Discussions (FGDs):
  ▪ A total of 12 FGDs will include participants such as: a) with male patients (2), b) female patients (2), c) community representatives (2), d) healthcare workers (1), e) local dispensers, traditional healers, and Traditional Birth Attendants (TBAs) (2), f) local NGO representatives (1) g) Family members of HCV-positive individuals (2). Each group will consist of 6 to 8 individuals.
- Key Informant Interviews (KIIs):
  ▪ Selected male patients (3)
  ▪ Selected female patients (3)
  ▪ Barbers (3)
  ▪ Hazam / traditional practitioners for circumcisions (4)
  ▪ Commercial sex workers (4)
  ▪ Selected community representatives (5)
  ▪ NGOs working on HIV and STI (3)
  ▪ Healthcare workers
    - Doctors (3)
    - Community Health Workers (2)
  ▪ Traditional healers (5)
  ▪ Dispensers (5)
  ▪ Family members of HCV-positive individuals (5)

Participants for FGDs and KIIs will be purposively sampled to ensure diversity in perspectives and experiences. This list could be readjusted, for instance, individuals from other categories could be included or more number of participants could be added in a given category, based on the findings from the quantitative study. The FGDs and KIIs will take place at camp level.

A semi-structured guiding questionnaire in English with translation in Rohingya language (Rohingyalish) will be used to guide the discussion for focus group discussion (FGD) and key informant interviews (KII). Only relevant questions related to the participant group will be asked. Similar to quantitative the questionnaire covers the risk factors in relation to socio-economic situation, medical history, traditional practices, high-risk behaviours and sexual practices. The research lead and trained research assistants will facilitate the FGDs and KII. FGD and KII will be conducted in the Rohingya language, and the responses will be translated in English in the report. An interpreter fluent in Rohingya language will be deployed for the FGDs and KII to eliminate the linguistic barriers. The average duration of each FGD will be around 90 minutes, and that of each KII will be 60 minutes.

### Selection Process

- FGDs: Participants will be selected based on their roles in the community such as healthcare workers, traditional healers, patients, or community representatives, and their relevance to factors influencing Hepatitis C transmission, including medical practices, behaviors, and exposure risks.
- KIIs: Informants will be chosen for their knowledge of specific practices or risk factors.

### Data Collection

- **Tool:** Semi-structured guiding questionnaires tailored for FGDs and KIIs, translated into Rohingyalish for cultural relevance.
- **Method:**
  ∘ FGDs will involve 6–8 participants per group and last ~90 minutes.
  ∘ KIIs will last ~60 minutes.
  ∘ Sessions will be audio-recorded with consent and transcribed for analysis.
- **Personnel:** Clinical supervisors, who are medical doctors with designated expertise, will oversee data collection to ensure quality and adherence to study protocols.

### Data Collection Instruments

#### Focus Group Discussion (FGD) Guide

A semi-structured guiding questionnaire (see SI1 File) will be used to conduct FGDs. The guide will be:

- **Language**: Developed in English and translated into Rohingyalish to ensure cultural relevance and participant comprehension.
- **Topics Covered:**
  ∘ Socio-economic factors (e.g., occupation, income, and education).
  ∘ Medical history (e.g., past surgeries, blood transfusions, and hospitalizations).
  ∘ Traditional practices (e.g., circumcision, piercing, and cupping).
  ∘ High-risk behaviors (e.g., sharing personal items, drug use).
  ∘ Sexual practices (e.g., unprotected sex, history of sexual violence).
- **Participant-Specific Questions:** Tailored questions will be included based on the participant group (e.g., male vs. female participants, healthcare workers).

#### Key Informant Interview (KII) Guide

The KII guide will include semi-structured questions to explore key insights from specific stakeholders. The guide will:

- **Language:** Follow the same format as FGDs, with English and Rohingyalish translations.
- **Topics Covered:**
  ∘ Detailed perspectives on identified risk factors (e.g., healthcare practices, hygiene standards).
  ∘ Community-level insights (e.g., stigma, healthcare access).
  ∘ Recommendations for targeted interventions.
- **Stakeholder-Specific Questions:** Tailored questions will address the expertise or experiences of the interviewees, such as:
  ∘ Barbers (practices around blade reuse).
  ∘ Traditional healers (cultural practices around circumcision or acupuncture).
- **Administration and Facilitation**
  ∘ Both FGDs and KIIs will be facilitated by trained research assistants under the supervision of the research lead.
  ∘ Sessions will be conducted in the Rohingya language to reduce linguistic barriers, with an interpreter fluent in Rohingyalish ensuring accuracy.
  ∘ Recording and Transcription: Sessions will be audio-recorded (with participant consent) and transcribed for analysis in English.
- **Session Duration**
  ∘ FGDs: ~90 minutes.
  ∘ KIIs: ~60 minutes.

### Data management and analysis

Quantitative data will be analyzed using Python, employing libraries such as Pandas, NumPy, SciPy, StatsModels, and scikit-learn. Socio-economic data and prevalence of the risk factors will be described in proportion and with mean, mode, median. Score of knowledge, attitude and practice will be derived by summing up each correct answer. Association of the risk factors to HCV positivity will be analyzed by simple and multiple logistic regression. Crude and adjusted odds ratios (ORs) will be presented with 95% CIs. Variables with a p-value of ≤0.2 in simple logistic regression analysis will be transferred into a multivariate regression model. Variables with a p-value of ≤0.05 in the multivariate regression model will be considered as significant risk factor.

The qualitative data, i.e., Key Informant Interviews (KII) and Focus Group Discussions (FGD), will be conducted in the Rohingya language and transcribed verbatim. The transcriptions will be translated into English for analysis. This approach involves identifying recurring themes, patterns, and insights from the qualitative data to gain a deeper understanding of the underlying factors and context. Data will be managed using NVivo software to facilitate coding and thematic extraction.

### Impact of research

∘ Assist in identifying key risk factors for Hepatitis C transmission among Rohingya population in camp settings in Cox’s Bazar, enabling targeted public health interventions and prevention strategies.
∘ Support healthcare providers in implementing safer medical procedures and infection control practices, improving health outcomes for vulnerable populations.
∘ Aid in designing effective community engagement and education campaigns by analyzing knowledge, attitudes, and practices related to Hepatitis C transmission.
∘ Provide evidence for policymakers to improve healthcare infrastructure and infection control measures in refugee settings, contributing to global strategies for addressing Hepatitis C in displaced populations.

## Ethical consideration

The research team received ethical clearance from the Ethical Review Board of Cox’s Bazar Medical College Hospital. The study protocol has also been reviewed and approved by the office of the Civil Surgeon and the Refugee Relief and Repatriation Commissioner (RRRC) Health Coordinator, who are responsible for overseeing activities related to the refugee crisis in Cox’s Bazar, Bangladesh. All necessary approvals have been obtained.

All participants will be asked to provide written informed consent, primarily through thumbprints. Consent documents will be securely stored separately from survey forms and transcripts. All surveys will be conducted by a trained enumerator of the same sex as the participant to ensure comfort and cultural sensitivity. Data protection measures will be strictly followed to maintain the confidentiality and privacy of all participants.

## Workplan

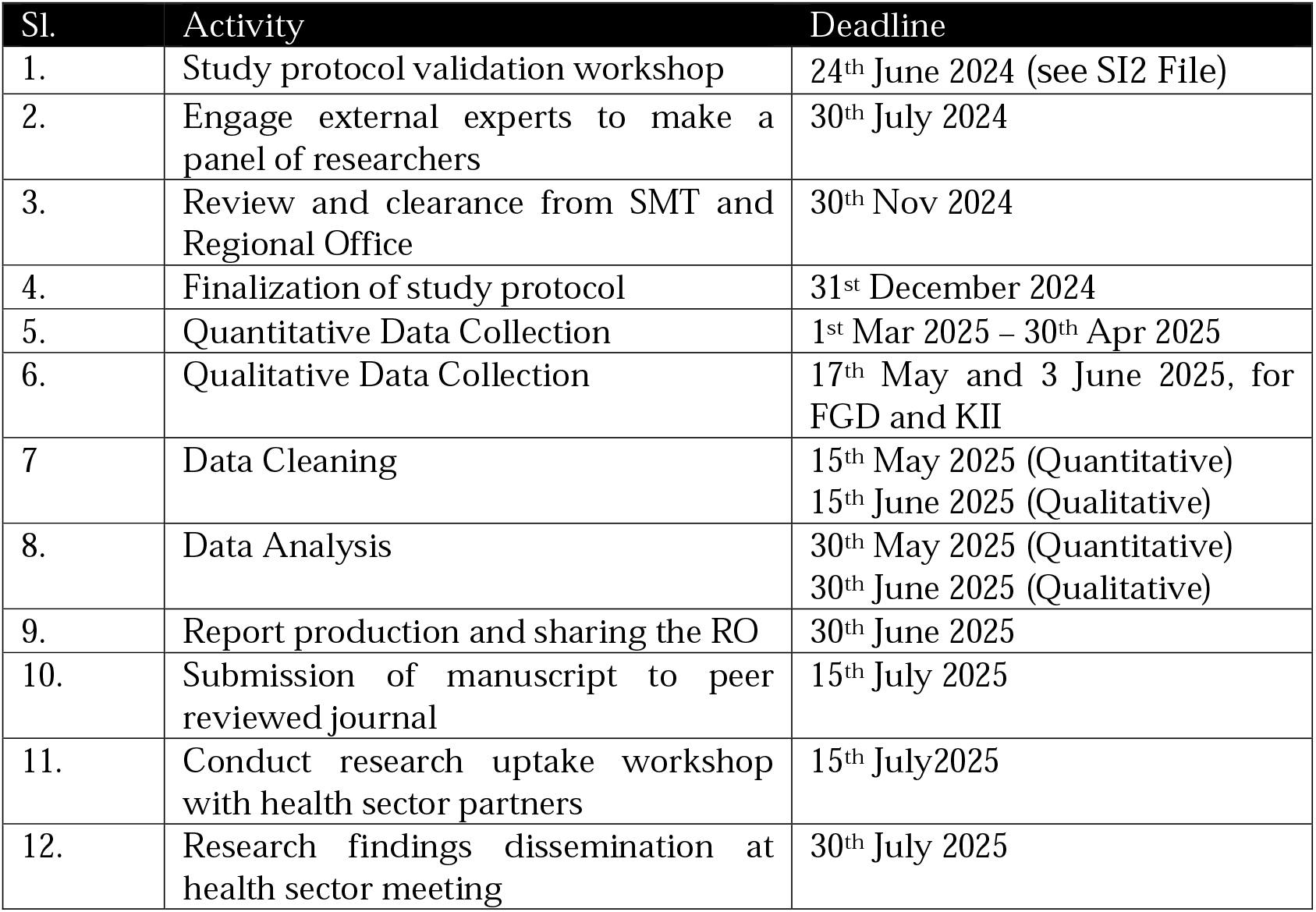

## Supporting information

SI1 File_Questionnaire (Semi-structured)

## Data Availability

All data generated or analyzed during this study will be made available upon reasonable request to the corresponding author.

## Supporting information

**S1 File**. Questionnaire (Semi-structured)

**S2 File**. Report of the Protocol Validation Workshop on Hepatitis C Research.

## Acknowledgments

This study will be conducted in the world’s largest refugee camps in Cox’s Bazar, Bangladesh. We express our sincere gratitude to all the experts, public health professionals, and clinicians who contributed to the review of this protocol.

