## Supplementary material for "Protocol for a Comprehensive Analysis of Hepatitis C Risk Factors among the Rohingya Population in Camp Settings in Cox’s Bazar: A Mixed-Method Study": SI1 File_Questionnaire (Semi-structured)

### QUANTITATIVE QUESTIONNAIRE:

**Introduction:**

*This survey is part of a research study to understand the risk factors associated with Hepatitis C transmission in the Rohingya refugee population. Participation is voluntary, and responses will remain confidential. Please answer honestly based on your personal experience. Some questions may not apply to you, and you may skip those as instructed.*

**Socio-Demographic:**

1. Age *(Numeric entry)*
2. Sex *(Male/Female)*
3. Type of refugee: ( Unregistered/ Registered)
4. Camp location:
5. Religion:
6. Born in: (Myanmar, Camps, elsewhere)
7. Education level: (No literacy, Can sign only, Below Grade 5, Grade 5 to 10, SSC equivalent, HSC equivalent, higher education)
8. Literacy status: Illiterate (unable to read or write), Semi-literate (able to sign or read basic texts), Literate
9. Marital status (married, single, divorced, widowed)
10. Occupation in Bangladesh:
11. Past occupation in Myanmar:
12. Arrival in Bangladesh (in years): *(Select: Before 2017, 2017, 2018, 2019, etc.)*
13. Number of family members:

**Instruction for Enumerators (**Questions 1-7:)**:** If the respondent has had multiple experiences for any of the following questions, ask them to base their responses on the **most recent incident**.

**Definition (Instruction/Guide for Enumerators)**

**Medical History:**

***Blood Transfusion:***

1. **Do you have a history of blood transfusion?** (Yes/No)
   a. **If yes:**
   b. **1.1 Where did it take place?**

*(Definitions:)*

   i. Myanmar: Within Myanmar before arriving at the camps.
   ii. Camps: Any medical facility inside the refugee camps.
   iii. Cox's Bazar: Hospitals or facilities outside the camps but in Cox's Bazar district.
   iv. Other: Any location not mentioned above (specify).

   c. **1.2 When did it take place?**

*(Options defined for time periods):*

i. Within last 5 years: Transfusion occurred in the recent past (up to 5 years ago).
ii. More than 5 - 10 years ago: Transfusion occurred between 5 and 10 years ago.
iii. More than 10 years ago: Transfusion occurred over a decade ago.

d. **1.3 What was the type of blood donor?**

*(Definitions for donor types):*

i. Family donor: Blood was donated by a family member.
ii. Voluntary non-remunerated donor: A donor who gave blood voluntarily without payment.
iii. Paid donor: A donor who was financially compensated.
iv. Donor from unknown source: The donor's identity or source is not known.

#### *Medical Injections:*

2. **Do you have a history of receiving medical injections?** (Yes/No)
   a. **If yes:**
      *i.* **2.1 Where did you receive the medical injections?**
      *(See location definitions in 1.1.)*
      *ii.* **2.2 When did it take place?**
      *(See time period definitions in 1.2.)*
      *iii.* **2.3 Who administered the injections?**
      *(Definitions for healthcare providers):*
         1. Healthcare workers at NGO/government health facility: Professionals working in recognized facilities.
         2. Local pharmacy/dispenser: Non-formal providers who administer injections.
         3. Traditional healers: Individuals using traditional or non-clinical methods.
         4. TBAs: Traditional Birth Attendants who provide injections during childbirth or related care.
         5. Others: Specify any provider not covered above.

      *iv.* **2.4 What type of syringe was used?**
      *(Definitions for syringe types):*
         1. Disposable syringe: A one-time use syringe.
         2. Reusable syringe: A syringe sterilized and used multiple times.
         3. Do not know: Respondent is unsure.

      v. **2.5 Have you ever been injected with a used needle?**
      (Yes/No/Do not know)

#### *Needle Prick Injuries:*

3. **Have you ever experienced a needle prick injury (e.g., from sharp objects like used needles) other than during medical injection?** (Yes/No)
   a. **If yes:**

*i.* **3.1 Where did it take place?**
*(See location definitions in 1.1.)*

*ii.* **3.2 How did the injury occur?**
*(Definitions for injury causes):*

1. Accidental handling of needle: Unintended handling of a used needle.
2. Handling sharp objects: Injury caused by contact with sharp materials.
3. Other: Specify any cause not listed above.

#### *Surgical Procedures:*

4. **Have you ever undergone a surgical procedure?** (Yes/No)
   a. **If yes:**
      *i.* **4.1 What type of surgery was it?**
      *(Definitions for surgery types):*
         1. Major surgery: Complex or invasive procedures requiring extended recovery.
         2. Minor surgery: Simple procedures with minimal recovery time.

      *ii.* **4.2 Where did the surgery take place?**
      *(See location definitions in 1.1.)*

      *iii.* **4.3 When did it take place?**
      *(See time period definitions in 1.2.)*

      **iv. 4.4 Who performed the surgery?**
      *(See provider definitions in 2.3.)*
      **[Instruction for Enumerator: Clarify that the aim is to assess who actually performed the procedure, irrespective of their authorization or qualification.]**

      v. **4.5 Have you ever been exposed to used and unsterile surgical instruments?** (Yes/No/Do not know)

      vi. **4.6 Have you ever been exposed to reused suture materials?** (Yes/No/Do not know)

#### *Hospitalization:*

5. **Have you ever been hospitalized?** (Yes/No)
   a. **If yes:**
      *i.* **5.1 Where were you hospitalized?**
      *(See location definitions in 1.1.)*

      ii. **5.2 Have you received intravenous injection or saline during hospitalization?** (Yes/No/Do not know)

#### *Dental Procedures:*

6. **Have you ever undergone any dental procedures?** (Yes/No)
    a. **If yes:**
        *i.* **6.1 Where did the procedure take place?**
        *(See location definitions in 1.1.)*
        *ii.* **6.2 Who performed the dental procedure?**
        *(Definitions for dental providers):*
            1. BMDC registered healthcare workers: Certified dental workers recognized by the Bangladesh Medical and Dental Council (BMDC).
            2. Local practitioner/dispenser: Informal or unregistered providers.
            3. Traditional healers/TBAs: See definitions in 2.3.
            *4.* Others: Specify any provider not listed above.
            *[Instruction for Enumerator: Licensed dental healthcare workers must be BMDC-certified.]
        iii. **6.3 Have you ever been exposed to used and unsterile dental instruments?** (Yes/No/Do not know)

#### *Hemodialysis:*

7. **Do you have a history of hemodialysis?** (Yes/No)
    a. **If yes:**
        *i.* **7.1 Where did the hemodialysis take place?**
        *(See location definitions in 1.1.)*

#### *Co-infections and Comorbidities:*

*8.* **Do you have any co-infections?**
*(Definitions):*
    a. HIV: Diagnosed through healthcare professionals with supporting medical records or tests.
    b. Hepatitis B: Diagnosed and confirmed through healthcare professionals or lab tests.
    c. Tuberculosis: Diagnosed and/or treated for TB in the past.

*9.* **Do you have any comorbidities?**
*(Definitions):*
    a. Diabetes Mellitus, Hypertension, COPD, Asthma, Ischemic heart disease, History of stroke, CKD, Mental health disorder (as diagnosed by healthcare professionals).

#### **Medical History:**

1. Do you have a history of blood transfusion? (Yes/No), If yes,
    - 1.1 Where did it take place? (Select all that apply: Myanmar, Camps, Cox's Bazar, Other: specify)
    - 1.2 When did it take place? (Within last 5 years, more than 5 - 10 years ago, more than 10 years ago)
    - 1.3 What was the type of blood donor? (Select one: Family donor, Voluntary non-remunerated donor, Paid donor, Donor from unknown source)
2. Do you have a history of receiving medical injections? (Yes/No). If yes,
    - 2.1 Where did you receive the medical injections? (Select all that apply: Myanmar, Camps, Cox's Bazar, Other: specify)
    - 2.2 When did it take place? (Within last 5 years, more than 5 - 10 years ago, more than 10 years ago)
    - 2.3 Who administered the injections? (Select all that apply: Healthcare workers at NGO/government health facility, Local pharmacy/dispenser, Traditional healers, TBAs, Others: please mention)
    - 2.4 What type of syringe was used? (Disposable syringe, Reusable syringe, Do not know)
    - 2.5 Have you ever been injected with a used needle? (Yes/No/Do not know)
3. Have you ever experienced a needle prick injury (e.g., from sharp objects like used needles) other than during medical injection? (Yes/No). If yes:
    - 3.1 Where did it take place? (Select all that apply: Health facility, Local pharmacy/dispenser, Own shelter, Other: specify)
    - 3.2 How did the injury occur? (Accidental handling of needle, Handling sharp objects, Other: specify)
4. Have you ever undergone a surgical procedure? (Yes/No). If yes,
    - 4.1 What type of surgery was it? (Major surgery, Minor surgery)
    - 4.2 Where did the surgery take place? (Select all that apply: Myanmar, Camps, Cox's Bazar, Other: specify)
    - 4.3 When did it take place? (Within last 5 years, more than 5 years ago)
    - 4.4 Who performed the surgery? (Select all that apply: Healthcare workers at NGO/government health facility, Local pharmacy/dispenser, Traditional healers, TBAs, Others: please mention)

[Instruction for Enumerator: Specify that this question aims to assess **who actually performed the surgery**, regardless of their authorization or qualification.]

    - 4.5 Have you ever been exposed to used and unsterile surgical instruments? (Yes/No/Do not know)
    - 4.6 Have you ever been exposed to reused suture materials? (Yes/No/Do not know)
    -
5. Have you ever been hospitalized? (Yes/No). If yes,
    - 5.1 Where were you hospitalized? (Select all that apply: Myanmar, Camps, Cox's Bazar, Other: specify)
    - 5.2 Have you received intravenous injection or saline during hospitalization? (Yes/No/Do not know)
6. Have you ever undergone any dental procedures? (Yes/No). If yes,
    - 6.1 Where did the procedure take place? (Select all that apply: Myanmar, Camps, Cox's Bazar, Other: specify)

- 6.2 Who performed the dental procedure? (Select all that apply: BMDC registered Healthcare workers at NGO/government health facility, Local practitioner/dispenser, Traditional healers, TBAs, Others: please mention)

[Note for enumerators: *Licensed dental healthcare worker:* Individuals formally trained and licensed to perform dental procedures from Bangladesh Medical and Dental Counsil (BMDC).]

- 6.3 Have you ever been exposed to used and unsterile dental instruments? (Yes/No/Do not know)

7. Do you have a history of hemodialysis? (Yes/No). If yes,
   - 7.1 Where did the hemodialysis take place? (Select all that apply: Myanmar, Camps, Cox's Bazar, Other: specify)
8. Do you have any co-infections:
   - HIV ((*documented: diagnosed by a healthcare professional and confirmed through medical records or laboratory tests* )
   - Hepatitis B (*diagnosed by a healthcare professional and confirmed through medical records or laboratory tests*)
   - Tuberculosis/ History of Tuberculosis (*diagnosed by a healthcare professional and/or treatment history*)
9. Do you have any comorbidities:
   - Diabetes Mellitus
   - Hypertension
   - COPD
   - Asthma
   - Ischemic heart disease
   - History of stroke
   - Chronic Kidney Disease (CKD)
   - Mental health disorder

(SKIP LOGIC: This question applies only to cases (individuals diagnosed with Hepatitis C).

10. Since being diagnosed with Hepatitis C, do you feel that your mental or emotional well-being has been affected? (Yes/No)

[Instruction: **For Enumerators:** Ensure that this question is skipped for participants in the control group; Verify that the participant understands the context of the question as related to their Hepatitis C diagnosis.]

11. Has your biological mother ever been diagnosed with HCV infection? (Yes/No/Do not know)
    - 11.1 If **Yes**, do you believe you were exposed to HCV through your mother during pregnancy, delivery, or breastfeeding? (Yes/No/Do not know)
    - 11.2 If **Yes** to Question 1, how many years before your diagnosis was your mother diagnosed? *(Please specify in years or select "Do not know")*

[Instruction for enumerators: Now I will ask about specific medical procedures related to childbirth or pregnancy. These questions are only for women who may have experienced complications during pregnancy or delivery. Please let me know if you are comfortable answering, and feel free to ask for clarification if needed.]

12. Have you undergone evacuation of retained products of conception? (Female only) (Yes/No). If yes,
    - 11.1 Where did the procedure take place? (Select all that apply: Myanmar, Camps, Cox's Bazar, Other: specify)
    - 11.2 Who performed the procedure? (Select all that apply: Midwife/skilled healthcare worker, Trained birth attendant, Traditional birth attendant)
    - 11.3 Were gloves used during the procedure? (Yes/No)
13. Have you ever experienced home delivery by a traditional birth attendant? (Yes/No). If yes,
    - 12.1 Where did the delivery take place? (Select all that apply: Myanmar, Camps)

**Traditional practices:**

14. Have you ever undergone acupuncture? (Yes/No)
15. Have you ever had any piercings (e.g., ear, nose)? (Yes/No). If yes,
    - 14.1 What type of piercing? Earring/nose ring/others (please mention)
    - 14.2 Was the piercing device/needle reused? (Yes/No/Do not know)
16. Do you have a history of cupping (bloodletting therapy)? (Yes/No)
17. Have you undergone circumcision by traditional or local practitioners outside of a hospital setting? (Male only) (Yes/No). If yes,
    - 16.1 Where did the circumcision take place? (Select all that apply: Myanmar, Camps, Cox's Bazar, Other: specify)
    - 16.2 Were you exposed to used and unsterile surgical instruments at that time? (Yes/No/Do not know)
    - 16.3 Who performed the circumcision? (Traditional practitioner/Hazom, Local dispenser/pharmacy practitioner, certified doctor, Other: specify)
18. Have you ever received a tattoo? (Yes/No)
19. Do you shave? (This question applies to male respondents only)
    - 19.1 How often? *(Often, Sometimes, Not usual)*
    - 19.2 Where do you shave? *(Home/Barber shop)*
    - 19.3 If at a barber shop, have you noticed if the barber uses a new blade or reuses a blade? *(New blade/Reused blade)*

**For both male and female:**

- 18.1 Have you used blades used by others?

For male only:

- 18.2 Where? (Home/Barber shop).
- 18.3 If barber shop, have you noticed if the barber changes the blade or use the same blade? (New blade/Same blade)

**Other high-risk factors**

20. Do you use recreational injection drugs? (Yes/No). If yes,
    - 18.1 Which drug do you use?
    - 18.2 Have you ever shared or reused someone else's needle? (Yes/No)
21. Have you ever been incarcerated? (Yes/No)
22. Does your spouse have HCV infection? (Yes/No/Not applicable)

23. Do you share any of the following personal items? (Razor, Toothbrush, Nail cutter) (Yes/No)

**Sexual activities/practices (sensitive questions, to be asked with cultural sensitivity)**

24. Have you had unprotected sex with an occasional partner? (Yes/No/Not willing to answer). If yes,
    22.1 How often? (Once only, Sometimes, Often)
25. Are you a man who has sex with men? (Yes/No/Not willing to answer) [Male respondents only]
26. Are you a woman working as a commercial sex worker? (Yes/No/Not willing to answer) [Female respondents only]
27. Do you get engaged in sexual intercourse in an unconventional way/route? (Yes/No/Not willing to answer)
28. Have you experienced sexual violence? (Yes/No). If yes,
    27.1 Where did it take place? (Select all that apply: Myanmar, Camps, Cox's Bazar, Other: specify)

**KAP survey:**

**Community Knowledge toward Hepatitis C Transmission and Prevention:**

1. Have you ever heard about a disease called Hepatitis C (HCV)? (yes/no)
    - If Yes: Proceed to questions 2–14.
    - If No: Skip the entire section and proceed to the next relevant section or end the survey (if applicable).
2. Is it caused by a virus? (yes/no)
3. Can Hepatitis C be transmitted by touching or holding hands with someone? (yes/no)
4. Is a Hepatitis C infected patient is at risk to others? (yes/no)
5. Is there any vaccine that can prevent Hepatitis C? (yes/no)
6. Does Hepatitis C affect the adolescents only? (yes/no)
7. Can Hepatitis C lead to liver cancer? (yes/no)
8. Can having injection with needle or syringe used by someone who has Hepatitis C spread the disease? (yes/no)
9. Can Hepatitis C spread through food or water? (yes/no)
10. Can hepatitis C spread through contaminated blood? (yes/no)
11. Can Hepatitis C be spread through unprotected sexual contact/intercourse? (yes/no)
12. Can Hepatitis C pass from mother to baby during pregnancy? (yes/no)
13. Can barbers using dirty blades spread Hepatitis C? (yes/no)
14. Can getting tattoos or piercings with unclean needles spread Hepatitis C? (yes/no)

**Community Attitude toward Hepatitis C Transmission and Prevention:**

15. Hepatitis C is a serious health problem. (Strongly Agree/ Agree/ Neutral/ Disagree/ Strongly Disagree)
16. People should get tested for Hepatitis C. (Strongly Agree/ Agree/ Neutral/ Disagree/ Strongly Disagree)

17. Having Hepatitis C affects a person’s ability to visit others. (Strongly Agree/ Agree/ Neutral/ Disagree/ Strongly Disagree)
18. It is safe to use the same cup or glass as someone who has Hepatitis C. (Strongly Agree/ Agree/ Neutral/ Disagree/ Strongly Disagree)
19. Injectable medicine and saline should only be received from a healthcare facility. (Strongly Agree/ Agree/ Neutral/ Disagree/ Strongly Disagree)
20. People who have multiple sexual partners are at a higher risk of getting Hepatitis C. (Strongly Agree/ Agree/ Neutral/ Disagree/ Strongly Disagree)

**Community Practices toward Hepatitis C Transmission and Prevention:**

21. When getting an injection, do you ask the medical staff to use a new, clean syringe? (Always/ Often/ Sometimes/ Rarely/ Never)
22. When getting a haircut or shave, do you ask the barber to use a new, clean blade? (might not be relevant for younger individuals who do not shave) (Always/ Often/ Sometimes/ Rarely/ Never)
23. Do you ask for new or sterilized instrument during circumcision of someone in your family? (might not be relevant for younger individuals) (Always/ Often/ Sometimes/ Rarely/ Never)
24. When getting a piercing or tattoo, do you make sure the needle is new and clean? (Always/ Often/ Sometimes/ Rarely/ Never)
25. Do you cover your open wounds and sores to prevent infection? (Always/ Often/ Sometimes/ Rarely/ Never)
26. Do you encourage people in your community to get vaccinated if a vaccine is available for Hepatitis C? (Always/ Often/ Sometimes/ Rarely/ Never)
27. If diagnosed with Hepatitis C, do you seek medical treatment promptly (Always/ Often/ Sometimes/ Rarely/ Never)

### QUALITATIVE QUESTIONNAIRE:

#### Introduction:

Thank you for participating in this discussion/interview. This study aims to explore your experiences and perspectives related to Hepatitis C and associated health practices within the Rohingya community. Your input is highly valuable and will contribute to designing effective strategies for prevention and management. Participation in this study is entirely voluntary and confidential. Please feel free to share your thoughts openly. If at any point you feel uncomfortable with a question, kindly let the researcher know, and you may choose not to respond.

#### Guidelines for FGDs and KIIs:

**Focus Group Discussions (FGDs)**:

- Target group: Community members (e.g., patients, caregivers, general population).
- Emphasis: Shared experiences, behaviors, and community norms.
- *Each FGD will cover 2-3 broad questions per theme to allow open, in-depth discussions.*

**Key Informant Interviews (KIIs):**

- Target group: Specific individuals with expertise or unique experiences (e.g., healthcare workers, TBAs, barbers, traditional healers, dispensers).
- Emphasis: Detailed insights on specific practices, observations, and expert opinions.

- 

**General:**

1. Can you describe the general living conditions for Rohingya refugees in the camps?
2. What are the common occupations or ways of earning a livelihood for refugees here?
3. Can you describe what kind of health care services are available in the camp?
4. In your opinion, are there any behaviors among this population that you think might increase the risk of hepatitis C transmission?

**Medical Exposure Related Questions:**

<u>Key Informant Interviews (KII)</u>

1. When do usually people have blood transfusions? Where does it usually take place? Share some your experiences from Myanmar and here in camp setting. Who performs the transfusions? Have you seen any reuse of blood transfusion appliances? If so, when and why these are reused? How can we prevent this?

2. Where do usually people receive medical injections both when you were in Myanmar and now in Bangladesh? Who administers these? Why do people prefer to use local pharmacies instead of health facilities? How the pharmacies get injectable products or injections or appliances? Are the devices or appliances reused? How and why are these reused? Is there any legal measures in place to prevent reuse of medical devices? How can we prevent this? Where are the syringes and needs disposed? Do the children and people get access to the used products?
3. Share your experiences on surgical procedures both when you were in Myanmar and now in Bangladesh? If people get injured, where do they seek care? Who do the surgeries? Why do people prefer to use alternative places other than NGO hospitals? What medical instruments are used for different surgeries? Are the instruments, devices or appliances reused? Do you know how these are sterilized? How can we prevent use of or exposure to unsterilized instruments. What is your experience about use of or exposure to used suture materials?
4. Share your experiences on dental procedures both when you were in Myanmar and now in Bangladesh. Who performs the dental procedures? Why do people prefer alternative places other than NGO hospitals? What medical instruments are used for different dental procedures? Are the instruments, devices, or appliances reused? Do you know how these are sterilized? How can we prevent exposure to unsterilized instruments?
5. Share your experiences in hospital settings. Is there any way people can get exposure to blood, blood products, or used medical devices and appliances?
6. Share your experience on hemodialysis both in Myanmar and Bangladesh.
7. What is your view about transmission of the disease from mother to child? Share your experience or from your neighbors or relatives.
8. Share your experience on home-based delivery. Why do people prefer home-based delivery instead of hospital-based delivery? How do TBAs influence childbirth? Do they use gloves? How does the Dai maintain sterility/cleanliness during home delivery?

Focus Group Discussions (FGD)

1. What traditional healing/treatment practices are commonly used among the refugees? Why you take this treatment? Do the traditional healers give any injections or do any suturing? Do they reuse the injection or suture? If yes, why?

2. Is there any other way people get needle prick injury from medical products? If so, usually where? Who are in most risk?

3. Do you think there is higher prevalence of Hepatitis C if people have comorbidity wither other condition (e.g. HIV, Hepatitis B, Diabetes, COPD, asthma, CKD, etc.?). If so, why?
4. Where do usually people receive medical injections in Myanmar and now in Bangladesh? How are these medical devices handled in the camp setting?
5. Are there common experiences regarding reuse of syringes or medical equipment among the community?

**Traditional practices:**

Key Informant Interviews (KII)

1. Share your experiences on the below traditional practices. What is the procedure? How is it related to your tradition and custom? Who performs? Where does it take place? What instruments are used and what is the source of instruments? Are the instruments reused? If so, are these sterilized? Why the instruments are reused and why are not sterilized?
    - Acupuncture (explain if the participant does not understand)
    - Piercing (ear, nose, etc)
    - Cupping (bloodletting therapy)
    - Circumcision*
    - Tattooing

    *Is there anyway health facilities can be accessed for circumcision? If no, why?

2. Share your experience on shaving practice for male and female? Where do people get the shaving instruments? Are these used by multiple individuals or family members? For male, in the barbar shop, what is the procedure of cleaning the device and changing the blades? Are the blades often reused? If so, why? How can we prevent reuse of the blades?

Focus Group Discussions (FGD)

1. Are there any other traditional practices, that exposes people to blood?
2. What are the community's thoughts and practices regarding shaving instruments shared among individuals or family members?

**Other high-risk factors:**

Key Informant Interviews (KII)

1. Share your experience on the use of recreational drugs by people in Myanmar and the camp settings. Who commonly takes these drugs? What kind of drugs are used?
2. Is there any social or legal system in place to prevent drug use?
3. How can we improve prevention of injectable drug use and needle sharing?

3.

Focus Group Discussions (FGD)

1. What are the community practices regarding sharing personal items such as razors, toothbrushes, and nail cutters?
2. Share any experiences of incarceration, particularly regarding sexual activities/practices and violence during prison stays.

**Sexual activities/practices:**

Key Informant Interviews (KII)

1. Share your views on transmission of Hepatitis C between spouses.
2. What are the attitudes toward contraception? What do you think about the use of protection (condoms) during sexual activity?

Focus Group Discussions (FGD)

1. Share your views on unprotected sex, men who have sex with men, commercial sex workers, unconventional sexual practices, and sexual violence.
    - Why do you think these practices are prevalent?
    - How can safe sexual practices be promoted?

**Knowledge, attitude and practice:**

Key Informant Interviews (KII)

1. What do you think about the Hepatitis C virus? How does it transmit?
2. Why do you think awareness about Hepatitis C remains low in the community?
3. What measures could help reduce the risk of Hepatitis C among the Rohingya refugees?
4. How can healthcare services be improved to meet the community's needs?

Focus Group Discussions (FGD)

1. Are there any stigmas associated with Hepatitis C or other infectious diseases in the refugee community?
2. Why do you think people lack a proactive attitude toward preventive measures?
3. What preventive measures do people follow, and what examples can you share?
